# Early outcomes of a large four-year MDMPH program: Results of a cross sectional survey of graduates (from three cohorts)

**DOI:** 10.1101/2022.09.04.22279585

**Authors:** Julia Belkowitz, Sabrina Payoute, Gauri Agarwal, Daniel Lichtstein, Roderick King, Shirin Shafazand, Latha Chandran

## Abstract

To address the public health needs of the 21^st^ century, the University of Miami Miller School of Medicine implemented a four-year MD/ MPH program in 2011 with a mission to graduate public health physician leaders. The first cohort of students graduated in 2015. In the summer of 2020, a survey was sent to graduates to gather information on their early career involvement in the areas of leadership, research, and public health. In addition to several multiple-choice questions, the survey included an open-ended question on the impact of public health training in their careers. Content analysis was used to analyze the responses to the open-ended question. Eighty-two graduates completed the survey; 80 of whom had participated in residency training. Forty-nine (61%) joined a residency in a primary care field. Many graduates had leadership roles in their early careers, including 35 (44%) who were selected as chief residents. Fifty-seven (71%) participated in research, most commonly in quality improvement (40, 50%), clinical (34, 43%) and community based (19, 24%). Over one third (30, 38%) chose to do work in public health during residency.

Themes that emerged regarding the impact of public health training on their careers were: 1) Shift in perspective, 2) Value of specific skills related to public health, 3) Steppingstone for professional opportunities 4) Focus on health disparities, social determinants, and inadequacies of the healthcare system, 5) Status as leaders and mentors for peers, and 6) Self-efficacy during the pandemic. Graduates self-reported significant commitment and involvement in leadership, research, and public health as well as towards addressing some of our most pressing public health needs. Although long-term career outcomes need to be determined over time, currently graduates report significant benefits of their public health training for their professional outcomes.

## Introduction

It is imperative that the US create a workforce that includes physicians trained in public health who have skills and competencies to address health issues beyond the individual patient to those involving the community and population. In 2007, the Institute of Medicine made a call to enhance public health training among physicians through multiple avenues, including formal degree programs[1]. The COVID-19 pandemic with its associated morbidity, mortality, and strain on our health system has highlighted the importance of social determinants of health and public health infrastructure in ensuring the health of our nation. Physicians who are trained in population and public health sciences play a valuable role in leading a responsive and transformative health system.

Over 80 medical schools currently offer opportunities for MPH training for their students, with most requiring a fifth year of training[2]. The number of programs has been increasing over the past decades, with a total of 822 graduates of combined MD/ MPH programs between 2007-2012 and 1,978 students who graduated between 2014-2019 [3,4].

MD/ MPH students self-reported greater levels of leadership, service and research as medical students as compared with MD only or other dual degree peers in a 2020 study reporting outcomes from a self-reported database of over 18,000 senior medical students [5]. Those evaluating career intentions and residency match outcomes comparing MD/ MPH program graduates with MD or other combined degree programs have found that MD/ MPH students are more likely to match in primary care specialties than peers [3,5,6] Krousel-Wood et al surveyed graduates who received public health training from one institution 10-20 years post-graduation and found that those with an MPH were more likely to be employed in academia, primary care, and/ or public health and were more likely to conduct research [7]. However, more detailed understanding of how graduates from other institutions focus their careers in public health, community service, research and leadership are needed, and no studies provide insight into the perceived impact on such training from the graduates themselves.

In 2011, with a goal of graduating public health physician leaders, the MD/MPH program at the University of Miami Miller School of Medicine (UMMSM) began enrolling yearly cohorts of about 50 students, thereby making it the largest such program in the country [4]. This cohort completes both degrees within the traditional four years of training in medical school through integration of both the medical school and Master of Public Health curricula. The program is designed to provide students with the academic knowledge and skills, as well as the clinical and public health experience, to improve the health of populations, especially those most vulnerable and underserved. The first cohort graduated in 2015, and there have been 328 MD/ MPH graduates of our program between 2015-2021. The purpose of this study is to describe the early career outcomes of the graduates of the UMMSM four-year MD/ MPH program in the graduating classes of 2015-2017 specifically related to their continued engagement in leadership roles, research, and public health after graduation.

## Methods

### Study design and participants

All graduates from the first three cohorts of the UMMSM MD/ MPH (UMM) program were eligible to participate in this cross-sectional survey. All eligible graduates for whom email addresses were available (130/142) were invited to participate in the online survey. The UMMSM Institutional Review Board approved this study (Protocol #20191104).

### Measures

Survey items were selected or adapted from surveys available through the American Association of Medical Colleges (AAMC) and other national organizations and finalized by this team of investigators[8,9,10,11,12].Demographic data was collected and included gender identity, marital status, number of dependents, and specialty of residency training as per the AAMC Graduation Questionnaire [8]. Race and ethnicity were defined according to the federal government definitions[9]. Types of residency training program was defined based on the AAMC Program Survey[10]. Graduates were also asked to identify age, graduation year and status of training in residency and fellowship (as applicable).

Questions about current workplace setting, were adapted from AAMC and American Academy of Pediatrics surveys [8,11,12]. Items regarding involvement in leadership, advocacy and public health were novel and included yes/ no or multiple responses with the opportunity for free text when graduates affirmed participation in a specific area (Table 1). The single open-ended question was “Please describe the impact of your public health education on your career.”

**Table 1.** Survey items for leadership, research, and public health.

| **Leadership** |
| --- |
| Were/ are you selected to be a chief resident? |
| Other than chief resident, did you/ do you hold other leadership positions as a resident? |
| Do you hold/ have you held leadership positions in your specialty organization? |
| Do you hold/ have you held other state or national leadership positions? |
| **Research** |
| Do you participate in research activities? |
| Have you applied for external funding (federal, foundation grants)? |
| Have you received any awards? |
| **Public Health** |
| Were you required to do any public health work during your residency training? |
| Outside of what was required, did you elect to do any public health work during your residency training? |
| What is the best description of your current work status? |
| What is the best description of your current place of practice? |
| Please indicate the setting in which you currently work. |
| Do you work in a primarily underserved area? |
| Please describe the impact of your public health education on your career. |

### Data analysis

Survey data was collected using Redcap v11.2.2, de-identified for analysis, and reported using tabular counts and descriptive statistics. Free text responses were independently summarized into categories by three investigators (JB, SP, GA) and the group discussed to achieve consensus when there was initial disagreement about the classification.

Qualitative analysis was done manually for the one open-ended question using conventional content analysis. Two investigators (JB and SP) independently reviewed the responses and together created a codebook summarizing the definition of the codes and explanation for how to apply the codes during the analysis. Three investigators (JB, SP and GA) then independently reviewed the free responses and applied the codes to the collected data. This team then consolidated the responses into broader categories and used those categories to identify themes that illustrated the perspectives of the participants about their experiences.

## Results

### Demographics characteristics

Of the 142 eligible participants, we had contact information on 130 alumni who were then invited to participate. This included 47/48 from the class of 2015 (98%), 45/48 from the class of 2016 (94%), and 38/46 (83%) from the class of 2017. A total of 82 alumni from the three cohorts responded to the questionnaire resulting in a 63% response rate. Demographic characteristics at enrollment in the survey are summarized in Table 2. Females represented 54% of survey respondents (44). Among survey respondents, 62% (51) self-identified as white, 4% (3) Black or African American, 13% (11) Asian, 11% (9) Hispanic, or Latino, or of Spanish origin, 6% multiracial (5), and 4% other race (3). The three cohorts were represented fairly evenly among the respondents. Seventy three percent (60) of respondents claimed zero dependents. Regarding marital status, 55% (45) of survey respondents were married. The average respondent age was 31.6 +/- 1.8 years (range 28 - 36 years).

**Table 2.** Demographic characteristics of participants.

|  | Total (n = 82) |
| --- | --- |
|  | Number (percent) |
| **Gender** |  |
| Female | 44 (53.7) |
| Male | 38 (46.3) |
| **Race and Ethnicity** |  |
| Asian | 11 (13.4) |
| Black or African American | 3 (3.7) |
| Hispanic, Latino, or of Spanish origin | 9 (11.0) |
| Multiple Race and Ethnicity | 5 (6.1) |
| White | 51 (62.2) |
| Other | 3 (3.7) |
| **Academic Year of Graduation** |  |
| 2015 | 31 (37.8) |
| 2016 | 29 (35.4) |
| 2017 | 22 (26.8) |
| **No. of Dependents (not including spouse/partner)** |  |
| 0 | 60 (73.2) |
| 1 | 14 (17.1) |
| > 1 | 8 (9.8) |
| **Marital Status** |  |
| Single (never legally married) | 32 (39.0) |
| Legally married | 45 (54.9) |
| Other | 5 (6.1) |
| **Age at time of survey, years** |  |
| Mean (SD): 31.6 (1.8) |  |
| Range: (28 - 36) |  |

### Residency and fellowship training

Of the total of 82 respondents, only 80 (98%) who participated in residency or fellowship training are included in the analysis. Of these, 35 participants had completed their graduate medical education. Details regarding status of training and specialty choice are depicted in Fig 1. Eighteen (23%) participated in a specialized track for residency training. Graduates in a specialized residency track reported to have participated in pathways such as Advocacy/ Community Health (5), Primary Care (4), Global Health (3), and others.

**Figure 1:**
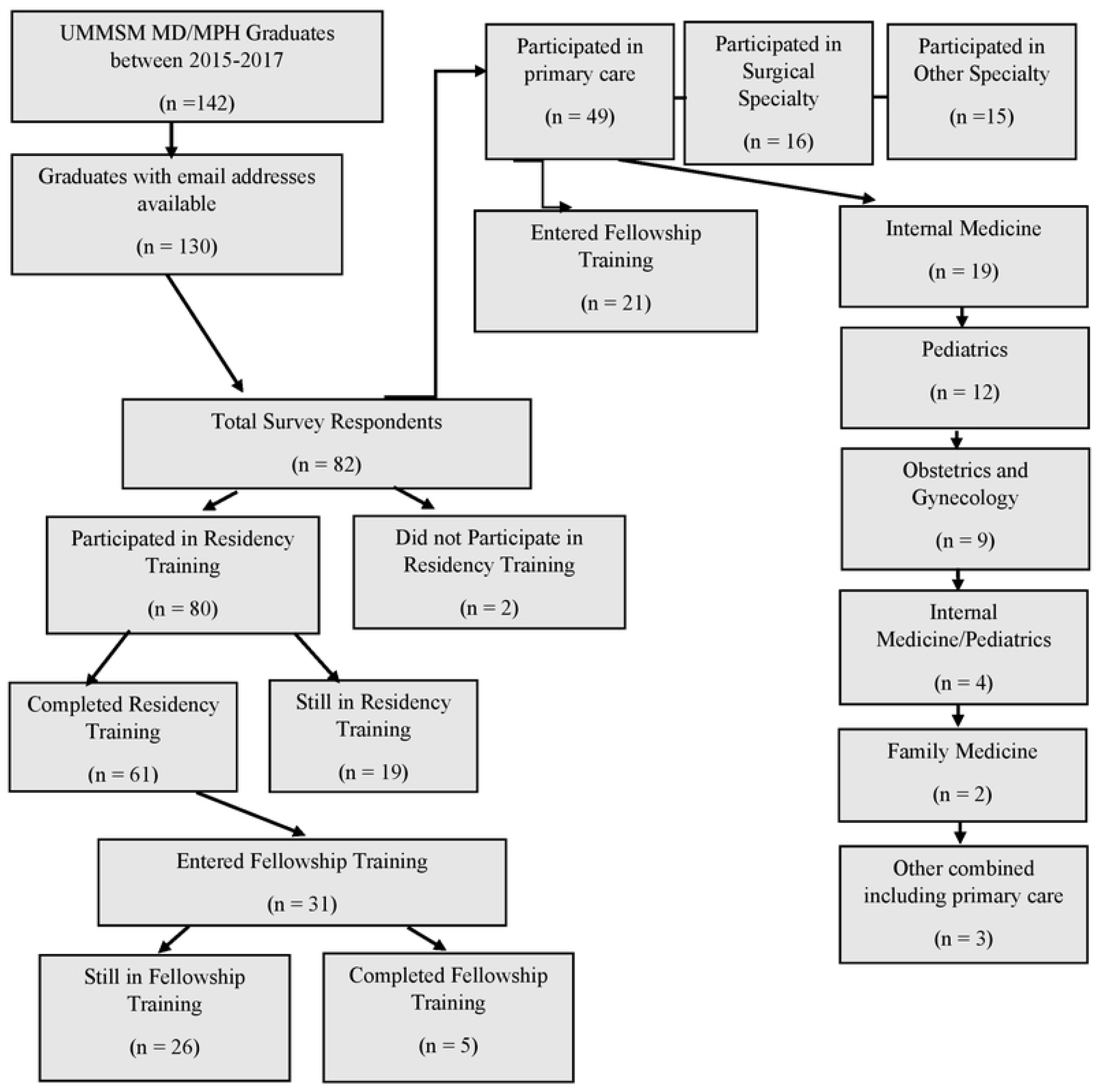
UMMSM MD/ MPH Alumni training status.

### Leadership, research, and public health

Many MD-MPH program graduates reported that they were appointed to a leadership position. Leadership characteristics are summarized in Table 3. Thirty-five (44%) were selected as chief residents. Thirty-eight (48%) listed other leadership activities held as a resident, 12 (15%) held leadership positions in their specialty specific organizations, and ten (12%) have held other state/national leadership positions. Categories of participation included leadership roles within their residency program or representing their residency program such as curriculum committee, Graduate Medical Education Committee representative (n=26); leadership within their specialty specific organization (ex. national delegate or committee chair), (n=18); leadership in research, patient safety, and/ or quality improvement (n=11); leadership regarding specific professional domains in the areas of diversity, wellness, and interprofessional collaboration (e.g. Diversity in Equity and Inclusion Committee), (n=10); leadership in community-based activities (e.g.. Community Health Committee) (n=5); and leadership related to clinical algorithms or skills development (eg. skills lab instructor) (n=4).

**Table 3:**
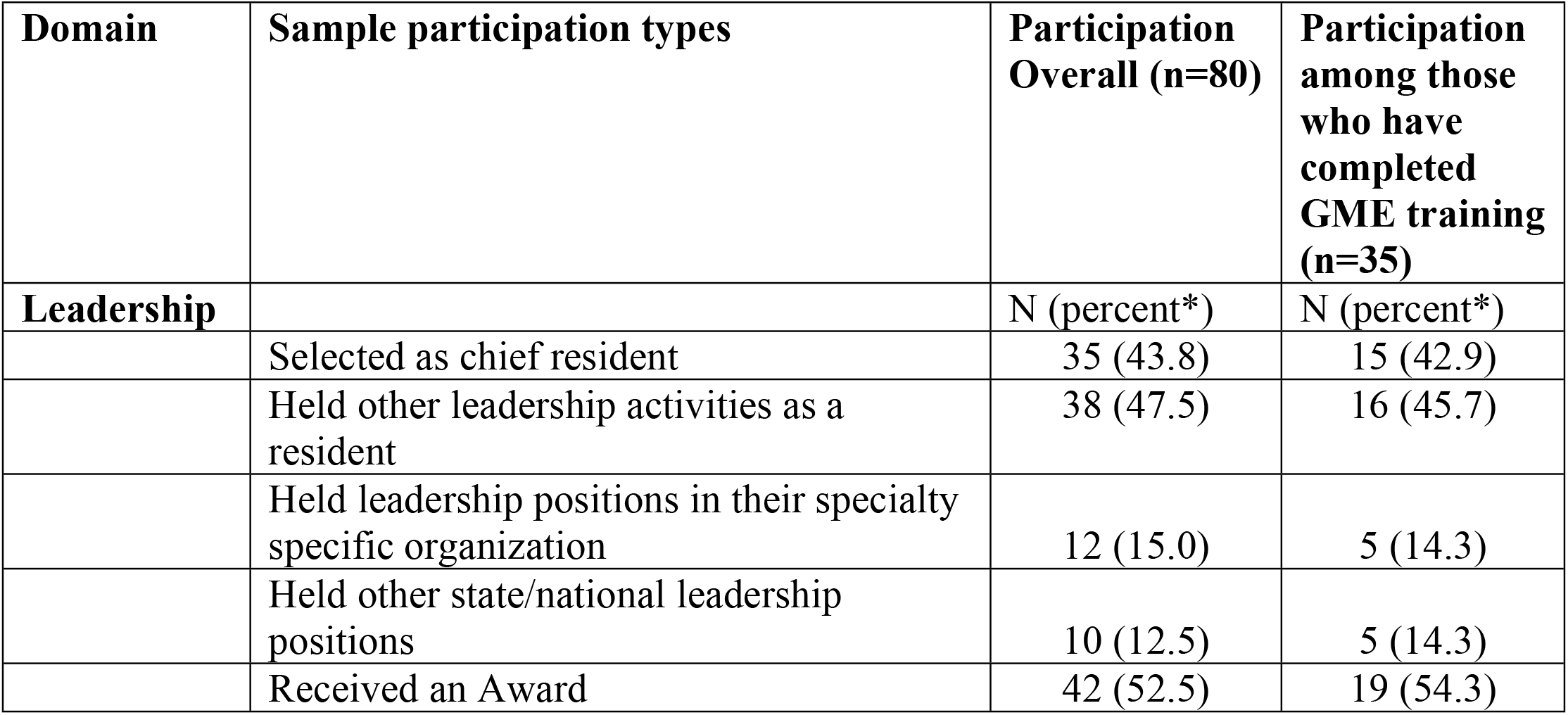
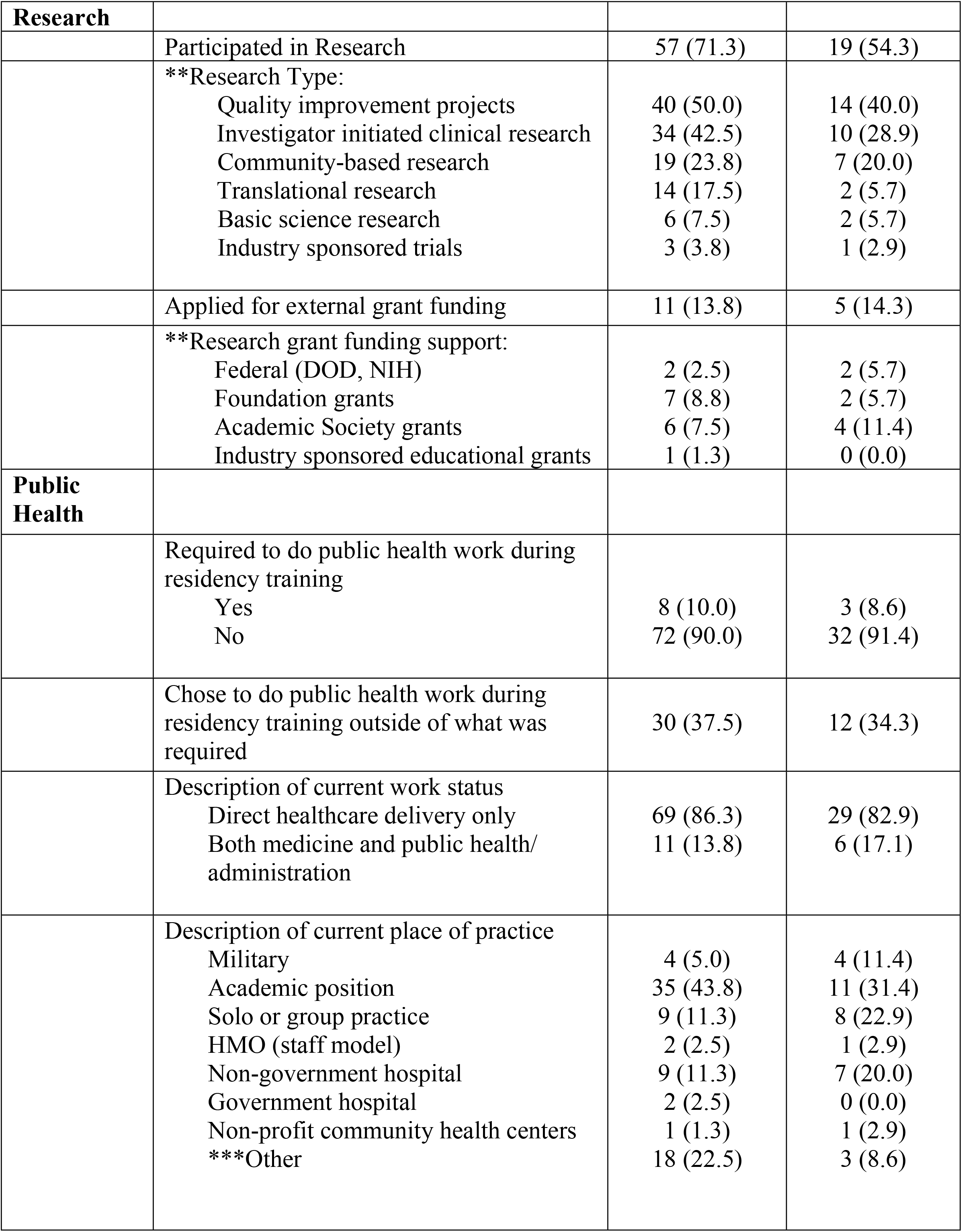
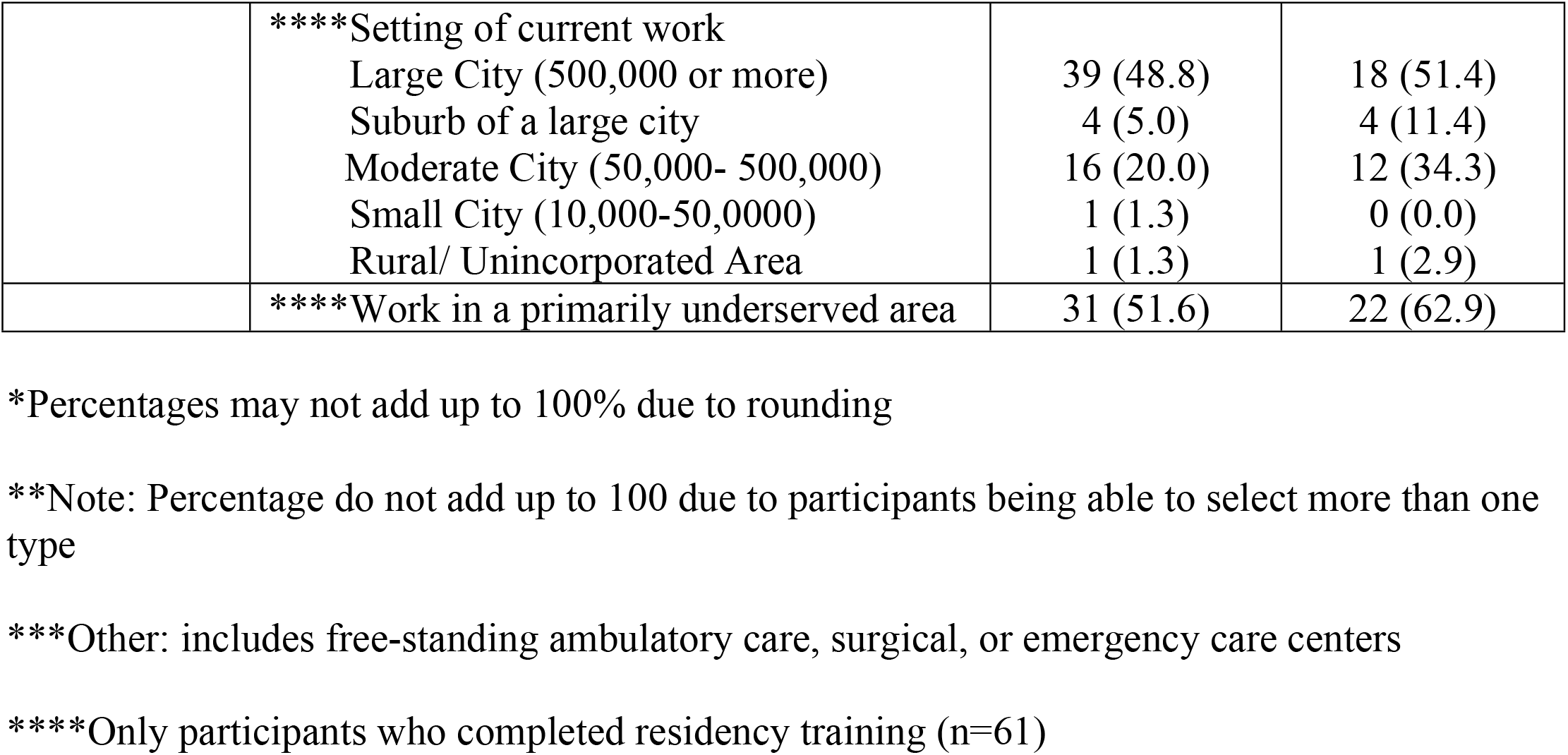
Leadership, research, and public health post medical school graduation.

| Domain | Sample participation types | Participation Overall (n=80) | Participation among those who have completed GME training (n=35) |
| --- | --- | --- | --- |
| **Leadership** |  | N (percent*) | N (percent*) |
|  | Selected as chief resident | 35 (43.8) | 15 (42.9) |
|  | Held other leadership activities as a resident | 38 (47.5) | 16 (45.7) |
|  | Held leadership positions in their specialty specific organization | 12 (15.0) | 5 (14.3) |
|  | Held other state/national leadership positions | 10 (12.5) | 5 (14.3) |
|  | Received an Award | 42 (52.5) | 19 (54.3) |
| **Research** |  |  |  |
|  | Participated in Research | 57 (71.3) | 19 (54.3) |
|  | **\*\*Research Type:**<br>Quality improvement projects<br>Investigator initiated clinical research<br>Community-based research<br>Translational research<br>Basic science research<br>Industry sponsored trials | 40 (50.0)<br>34 (42.5)<br>19 (23.8)<br>14 (17.5)<br>6 (7.5)<br>3 (3.8) | 14 (40.0)<br>10 (28.9)<br>7 (20.0)<br>2 (5.7)<br>2 (5.7)<br>1 (2.9) |
|  | Applied for external grant funding | 11 (13.8) | 5 (14.3) |
|  | **\*\*Research grant funding support:**<br>Federal (DOD, NIH)<br>Foundation grants<br>Academic Society grants<br>Industry sponsored educational grants | 2 (2.5)<br>7 (8.8)<br>6 (7.5)<br>1 (1.3) | 2 (5.7)<br>2 (5.7)<br>4 (11.4)<br>0 (0.0) |
| **Public Health** |  |  |  |
|  | Required to do public health work during residency training<br>Yes<br>No | 8 (10.0)<br>72 (90.0) | 3 (8.6)<br>32 (91.4) |
|  | Chose to do public health work during residency training outside of what was required | 30 (37.5) | 12 (34.3) |
|  | Description of current work status<br>Direct healthcare delivery only<br>Both medicine and public health/ administration | 69 (86.3)<br>11 (13.8) | 29 (82.9)<br>6 (17.1) |
|  | Description of current place of practice<br>Military<br>Academic position<br>Solo or group practice<br>HMO (staff model)<br>Non-government hospital<br>Government hospital<br>Non-profit community health centers<br>***Other | 4 (5.0)<br>35 (43.8)<br>9 (11.3)<br>2 (2.5)<br>9 (11.3)<br>2 (2.5)<br>1 (1.3)<br>18 (22.5) | 4 (11.4)<br>11 (31.4)<br>8 (22.9)<br>1 (2.9)<br>7 (20.0)<br>0 (0.0)<br>1 (2.9)<br>3 (8.6) |
|  | ****Setting of current work |  |  |
|  | Large City (500,000 or more) | 39 (48.8) | 18 (51.4) |
|  | Suburb of a large city | 4 (5.0) | 4 (11.4) |
|  | Moderate City (50,000- 500,000) | 16 (20.0) | 12 (34.3) |
|  | Small City (10,000-50,000) | 1 (1.3) | 0 (0.0) |
|  | Rural/ Unincorporated Area | 1 (1.3) | 1 (2.9) |
|  | ****Work in a primarily underserved area | 31 (51.6) | 22 (62.9) |

\*Percentages may not add up to 100% due to rounding
\*\*Note: Percentage do not add up to 100 due to participants being able to select more than one type
\*\*\*Other: includes free-standing ambulatory care, surgical, or emergency care centers
\*\*\*\*Only participants who completed residency training (n=61)

Over half of those who had undergone residency training (n= 42, 53%) reported receiving an award professionally. Categories included awards highlighting their excellence as a resident/clinical skills from either the residency program or the specialty society (n=33); awards for research/ scholarship or patient safety/ quality improvement (n=21); awards for teaching/ humanism/ AOA (n=16); and other awards (n=12).

Fifty-seven (71%) MD-MPH program alumni reported participation in research. Research characteristics are summarized in Table 3. Forty (50%) participated in quality improvement projects, 34 (43%), participated in investigator initiated clinical research, 19 (24%) in community-based research, 14 (18%) in translational research, 6 (8%) in basic science research, 3 (4%) in industry sponsored trials. Among the 57 that participated in research, 11 (14%) successfully received research grant support from sources including federal agencies (2, 3%), foundation (7, 9%), academic society (6, 8%), and industry sponsored grant funding (1, 1%).

Results regarding the graduates’ involvement in public health are summarized in Table 3. Over a third of the respondents, 30 (38%) voluntarily chose to participate in public health work during residency, outside of what was required of them. Overall, categories of the public health related work were included Advocacy/ leadership (n=14), Research/ patient safety/ quality improvement (n=12), community based (n=10), global health (n=8), clinically related (n=6), teaching (n=5).

Analysis of the responses to the question to “describe the impact of your public health education on your career” revealed six broad themes: 1) Shift in perspective, 2) Value of specific skills related to public health, 3) Stepping stone for professional opportunities 4) Focus on health disparities, social determinants and inadequacies of the healthcare system, 5) Status as leaders and mentors for peers, and 6) Self-efficacy during the pandemic. Further details are described in Table 4.

**Table 4:** Qualitative analysis results for impact of public health training.

| Theme | Description | Representative Quotation(s) |
| --- | --- | --- |
| Shift in Perspective | Overall career and approach to clinical medicine. | • “My public health education has shaped the way I practice medicine in many ways. Every time I see a patient I try and get to the root cause of their problem(s),”<br>• “Public health has formed the way I view medicine. My perspective is different because of my public health training. I view things from a systemic perspective in addition to an individual patient perspective, and try to be cognizant of systemic issues which prevent adequate health in the community in which I practice.” |
|  | Direct impact on their personal interactions with patients | • “Changes the way I treat patients. Helps me connect on a deeper level with people from all backgrounds,” |
|  | Related to their own lives | • “My public health education taught me to better understand why underserved populations have worse health outcomes, which has inspired me to actively work to eliminate this health outcome gap by working on implicit personal level and system level biases in the way I practice medicine and live my life.” |
| Value of specific skills related to public health | Content from the formal public health curriculum as important for their current day to day work. | • “My [public health] education has given me the ability to easily critically appraise medical literature and understand the meaning of biostatistics”<br>• “The biggest impact I have found is how my training has helped me interact with hospital administration to improve patient care in both the inpatient and outpatient center.” |
|  | Capability with research in a variety of domains | • “The coursework of the MPH degree has proven useful in clinical research, project proposals, grant applications, and publication in peer-reviewed journal”<br>• “I gained substantial research and statistical training through my public health education that I continue to apply to my current research in high-risk women and infectious disease”<br>• “My public health background has encouraged me to push for or find a solution to increase minority/ underserved population inclusion in cancer clinical trials. I initiated a retrospective study based on rising costs of [disease management] to find targeted areas to help with this issue.” |
| Stepping stone for professional opportunities | Workload demands limited their opportunity to fully participate in public health | • “While much of residency has been taken up with clinical duties, I’m still hoping to pursue a research focus in healthcare disparities as an attending.” |
|  | Access and meaningful contribution to positions | • “My training also made me better suited to function as a leader and community liaison for the [community-based program].”<br>• “My public health education has been incredibly important for my career. It has allowed me to understand public health and to pursue [professional opportunity] which has been an amazing experience.” |
| Focus on health disparities, social determinants and inadequacies of the healthcare system. | Integration of social determinants into daily work and approach to healthcare | • “It created a strong foundation of the importance of population health and understanding how social determinants have a huge impact on individuals health and further more on communities”<br>• “Aspects of my clinical research and quality improvement projects and seek to improve healthcare systems and delivery of care within our [clinic]. I feel strongly that it is my public health background that has provided the skills and knowledge to carry all of this out...”<br>• “Exposed me to working collaboratively and leading groups around population- specific priorities and opportunities for improvement in healthcare delivery and health promotion.” |
|  | Working with vulnerable populations | • “I work at [county’s] public safety net hospital and am committed to a career in service to underserved populations.” |
| Status as leaders and mentors for peers | Viewed as leaders in their residency/ work because of their specialized training | • “Set me apart from my peers as being uniquely qualified in healthcare systems- thinking and related leadership roles”<br>• “My co-residents often come to me for help with their quality improvement projects and |
|  |  | questions regarding interpreting the medical literature because they know that I have/had public health and research training.” |
|  | Teaching was identified as an important aspect of this status | • “...Also, as a public health physician I try and educate my colleagues and patients on the importance of public health.” |
|  | Advocates for their patient population | • “...it pushed me to strength my skills in advocating for my patient and finding the right resources for them.” |
| Self-efficacy during the pandemic | Felt especially equipped to be working in the healthcare system as the COVID-19 pandemic emerged. | • “It has also helped me to understand the COVID pandemic at the epidemiologic and socio-political levels, even from very early on.” |

## Discussion

This study is the first to examine early career outcomes and qualitative impact from a large cohort of MD/ MPH students. Our study results indicate that the graduates of the UMMSM dual degree MD/MPH program are utilizing their public health education and continue to remain committed to leadership, research, and public health several years after graduation. The graduates’ choices of post-graduate programs (residencies) indicated that over 50% of them chose primary care specialties or combined primary programs that included primary care. Though the definition of primary care varies in the literature, this team of investigators elected to define primary care to include family medicine, internal medicine, pediatrics and obstetrics and gynecology, consistent with the United States Health Resources & Service Administration definitions [7,13,14,15]. The tendency towards MD/ MPH graduates to more heavily choose primary care residencies is consistent with graduate intentions and match outcomes at other institutions [3,5,6]. In a long-term study which followed graduates from one institution’s MD/ MPH program for 10-20 years post-graduation, members of their MD/ MPH cohort were more likely to practice general primary care [7]. It remains to be seen if our graduates will indeed follow a similar path as, 21 (43%), of our graduates who initially entered primary care related residencies ultimately participated in fellowship training. Future studies investigating the long-term career pathways will be required to better understand their ultimate career trajectories.

Nonetheless, the mission of the dual degree program at our institution is not rooted in training primary care physicians. Rather, the stated mission is to develop public health physician leaders, regardless of specialty choice [16]. The results of our study indicate that they are indeed beginning to build a foundation to become public health leaders based upon their described involvement in leadership and research experiences. Forty four percent of the program’s alumni were selected as chief residents, and 48% held some other leadership role as a resident outside of serving as chief. Seventy one percent of the graduates participated in research since graduation, with 50% involved in quality improvement related work and 24% in community-based research. Narrative comments also supported their trajectory towards becoming public health leaders through their self-identified positions in their residency program as role models and teachers. Graduates of other MD/ MPH programs are more likely to describe intentions of working in academic medicine, healthcare administration and/or state and federal administration [3]. Christensten et al described residency characteristics comparing MD/ MPH students with other dual degree or MD only graduates and found that MD/ MPH students had significantly more participation in Gold Humanism Honor Society membership, research related experiences, leadership experiences, volunteer experiences during medical school [5]. We intend to follow this and future cohorts over time to better describe the ultimate career outcomes of our graduates.

An important question is whether public health training in medical schools will lead to an expansion of the workforce of public health professionals. Emerging findings from this study and others seem to support that it will. More than one third (38%) of the graduates voluntarily elected to participate in public health endeavors during residency above and beyond what was required in their respective curricula, despite the rigorous demands of their schedules and residency responsibilities. Perhaps most meaningful in evaluating the impact of public health education (MPH) on the careers of the respondents were the themes that emerged from the rich narrative comments about the impact of the program’s public health training on their careers. Not surprisingly, students appreciated the specific public health skills such as appraisal of the literature and research. Others have surveyed medical student participants of their MPH training programs and similarly reported improved skills in critical appraisal of the literature and research skills, as well as a commitment towards underserved populations, policy and global health [13,17]. A national study of a cohort of 822 MD MPH graduates found that MD MPH program graduates were more likely to report altruism as a factor in their career choice than MD only graduates [3]. These findings are also highlighted in the comments from our study participants in themes that emerged regarding dedication towards addressing social determinants and service towards vulnerable populations, among others.

Limitations include that this data was self-reported using a survey that had not been formally validated. There is always a potential for self-selection bias given the methodology. It is also difficult to make predictions regarding the long-term involvement of study participants in leadership, research, and public health, and to determine this we plan to follow this and future cohort of our graduates to determine the level of their continued engagement these areas.

This study demonstrates that integrating a Master of Public Health curriculum into a four-year combined medical school program can lead to the successful development of a cohort of dedicated public health physician leaders. Though the full impact of this training cannot be measured for many years to come, it is encouraging to see the early career outcomes of MD/MPH trained physicians. Especially given the burgeoning public health crises resulting from the current pandemic, climate change, and other health needs around the world, it is the responsibility of the medical education community to continue to expand similar training programs that integrate the public health work force needs with the idealism and altruism that many students bring to their training.

## Data Availability

The survey data was collected using RedCap and downloads are stored in our institution's Box folders.

## Acknowledgments

The authors wish to thank Michael Maguire, MD, MPH for his support in providing contact information for alumni, Marta Bergez for providing data regarding the MD/ MPH cohort, and Anna Yabloch for her support in initial technical management of the survey.

## Notes

### Competing Interest Statement

The authors have declared no competing interest.

### Funding Statement

The author(s) received no specific funding for this work.

### Author Declarations

The University of Miami Miller School of Medicine Institutional Review Board approved this study (Protocol #20191104).

